# Impact of Daily Physical Fatigue on Inter-Limb Load Asymmetry and Joint Loading Rates During Daily Living Tasks in Unilateral Knee Osteoarthritis

**DOI:** 10.64898/2026.09.25.26364015

**Authors:** Muhammad Hassan Bari, Ghulam Abbas, Abdullah Abid, Atta Rasool

## Abstract

**Background:** Knee osteoarthritis (OA) is characterized by progressive joint degradation and altered gait biomechanics. Inter-limb load asymmetry during daily living tasks may accelerate joint deterioration, particularly when combined with physical fatigue. This study investigates the relationship between daily physical fatigue and inter-limb load asymmetry patterns during functional activities in individuals with unilateral knee OA.

**Objective:** To evaluate the impact of fatigue on inter-limb load asymmetry and joint loading rates during daily living tasks in patients with unilateral knee osteoarthritis, and to identify biomechanical and clinical factors associated with load distribution patterns.

**Methods:** One hundred eighteen participants with unilateral knee OA (mean age 62.6 (SD 7.4) years, BMI 29.9 (SD 4.2) kg/m^2^) completed biomechanical assessments during non-fatigued and fatigued conditions. Ground reaction force measurements were obtained during functional tasks including stair climbing, sit-to-stand transitions, and level walking. Load asymmetry was quantified as the inter-limb difference in peak vertical ground reaction forces. Statistical analysis was conducted using Python (v3.9), including paired t-tests, analysis of variance, and linear regression modeling.

**Results:** Participants demonstrated significantly increased load asymmetry under fatigue conditions (mean difference 8.7% (SD 12.3) %, *p* < 0.001). Fatigue-induced asymmetry was task-dependent, with stair negotiation showing the greatest increase (11.2% (SD 14.1) %). Peak loading rates on the affected knee were inversely correlated with time-to-peak load during fatigued conditions (*r* = -0.68, p < 0.001). Disease severity (WOMAC score) and age significantly predicted asymmetry changes (R^2^ = 0.52, *p* < 0.001).

**Conclusions:** Physical fatigue exacerbates inter-limb load asymmetry in unilateral knee OA, potentially accelerating joint degeneration. Task-specific interventions targeting fatigue management and load redistribution may improve long-term outcomes. Future longitudinal studies should examine the temporal relationship between acute biomechanical changes and progressive structural joint damage.

## Introduction

Knee osteoarthritis (OA) represents one of the most prevalent musculoskeletal disorders affecting the elderly population, with an estimated prevalence of 28% in individuals over 45 years of age globally (1). The condition is characterized by progressive cartilage degeneration, subchondral bone remodeling, and inflammatory processes that substantially impair functional capacity and quality of life (2). While genetic predisposition, age, and obesity constitute well-established risk factors, emerging evidence suggests that biomechanical factors, particularly load distribution patterns, play a critical role in disease progression (3, 4).

Inter-limb load asymmetry during gait and functional activities has been extensively documented in OA populations, where the affected limb typically experiences reduced load-bearing to minimize pain and joint stress (5). However, this compensatory mechanism paradoxically perpetuates asymmetry by facilitating muscular deconditioning of the affected limb and imposing disproportionate loads on the contralateral limb (6). Longitudinal biomechanical studies have demonstrated that persistent asymmetrical loading patterns are associated with accelerated joint space narrowing and radiographic progression of OA (4, 7).

Physical fatigue represents a significant yet understudied factor that may modify load distribution during functional activities. Fatigue-induced neuromuscular changes, including reduced muscle activation amplitude, delayed motor recruitment, and impaired proprioceptive feedback, are known to alter gait kinematics and kinetics (8, 9). In individuals with knee OA, fatigue-related alterations in muscle activation may exacerbate compensatory gait strategies and interlimb asymmetries, potentially increasing mechanical loading on the contralateral limb and contributing to further joint degeneration (10). However, empirical evidence quantifying the impact of fatigue on inter-limb load asymmetry in OA populations remains limited.

Daily living tasks, including stair negotiation, sit-to-stand transitions, and level walking, represent high-demand functional activities that generate substantial joint forces (11). The peak vertical ground reaction forces during these activities can exceed 2.0-3.0 times body weight, particularly during stair descent (12). Individuals with unilateral knee OA likely adopt task-specific compensatory strategies; therefore, the relationship between fatigue and load asymmetry may be task-dependent (13). Understanding these relationships is essential for developing targeted interventions that mitigate fatigue-related biomechanical deterioration.

Fatigue-related gait alterations are not limited to load asymmetry alone: spatiotemporal changes such as reduced gait speed and shortened stride length have been documented in older adults with a history of falls (14, 15), and sex-specific differences in fatigue-induced gait adaptation have been reported in populations with anterior knee pain (16). External factors such as footwear characteristics can further modulate gait patterns in knee OA, independent of body weight (17), underscoring the multifactorial nature of load regulation in this population. Reliable quantification of disease burden alongside these biomechanical measures typically relies on validated patient-reported outcome instruments such as the WOMAC index (18).

The primary objectives of this study were:

1. To quantify changes in inter-limb load asymmetry under fatigued versus non-fatigued conditions.
2. To evaluate task-specific variability in fatigue-induced asymmetry patterns.
3. To determine clinical and demographic factors predicting asymmetry responses to fatigue.
4. To investigate correlations between loading parameters and disease severity.

We hypothesized that physical fatigue would significantly increase inter-limb load asymmetry, with pronounced effects during high-demand tasks such as stair negotiation.

## Methods

### Study Design and Participant Recruitment

This was a cross-sectional observational study incorporating a repeated-measures design comparing biomechanical parameters during non-fatigued and fatigued conditions. Participants were recruited from the outpatient clinics at Islam College of Physical Therapy and affiliated orthopaedic centers in Sialkot, Pakistan, between March 2024 and December 2025. The study followed the ethical principles of the Declaration of Helsinki and was approved by the Institutional Review Board of Islam College of Physical Therapy, all participants provided written informed consent prior to enrolment.

### Participant Selection Criteria

#### Inclusion criteria

1. Age ≥ 45 years.
2. Radiographically confirmed unilateral knee OA (Kellgren-Lawrence grade II-IV).
3. Symptom duration ≥ 6 months.
4. Ability to ambulate independently without assistive devices.
5. Ability to perform functional tasks (stair climbing, sit-to-stand transitions).
6. No serious medical comorbidities precluding exercise participation.

#### Exclusion criteria

1. Bilateral knee OA.
2. History of knee arthroplasty or arthroscopic surgery within the previous 12 months.
3. Acute knee effusion or inflammatory conditions.
4. Neurological disorders affecting motor control.
5. Lower limb injuries unrelated to OA within the previous 6 months.
6. Cardiovascular contraindications to exercise testing.
7. Use of analgesic or anti-inflammatory medications within 48 hours of assessment.

### Clinical Assessments

Participant demographics including age, sex, body mass index (BMI), disease duration, and affected limb were recorded. Disease severity was quantified using the Western Ontario and McMaster Universities Osteoarthritis Index (WOMAC), a validated 24-item questionnaire assessing pain (5 items), stiffness (2 items), and physical function (17 items), with total scores ranging from 0 to 96 (18).

### Biomechanical Assessment Protocol

Participants performed three standardized functional tasks:

a. Level walking at self-selected pace over a 10-meter walkway.
b. Sit-to-stand transitions from a standard height chair (45 cm).
c. Stair negotiation (ascending and descending a 4-step staircase with standard dimensions: 17 cm rise, 28 cm tread depth).

Kinetic data were collected using two synchronized force plates (Bertec Corporation, Columbus, OH) integrated into a biomechanical analysis system with a sampling frequency of 1000 Hz. Participants performed three trials of each task under two conditions:

i. Non-fatigued condition, performed at the beginning of the assessment session following a 10-minute rest period.
ii. Fatigued condition, performed following a standardized fatigue protocol consisting of 6 minutes of continuous stepping at 60 steps/minute (equivalent to approximately 360 total steps), followed immediately by the functional tasks without rest.

Fatigue was confirmed using a modified Borg Rating of Perceived Exertion scale (0–10 scale) (19), with the criterion of ≥ 7/10 perceived exertion required to proceed with fatigued-condition testing.

### Data Analysis Using Python

Data processing and statistical analysis were conducted using Python version 3.9.7 with the following libraries: pandas (v1.3.5) for data manipulation, NumPy (v1.21.0) for numerical computations, SciPy (v1.7.0) for statistical testing, and Matplotlib (v3.4.3) for visualization.

Raw kinetic data were filtered using a fourth-order Butterworth low-pass filter with a cut-off frequency of 25 Hz (20). Peak vertical ground reaction forces were extracted for the affected and unaffected limbs during each trial. Inter-limb load asymmetry was quantified as the absolute percentage difference between peak vertical ground reaction forces normalized to body weight: **Asymmetry Index (%)** = |Peak Load (Affected) − Peak Load (Unaffected)| / [(Peak Load (Affected) + Peak Load (Unaffected))/2] × 100.

Summary statistics including means, standard deviations, and 95% confidence intervals were calculated for all biomechanical variables. Paired t-tests compared asymmetry indices between non-fatigued and fatigued conditions. One-way analysis of variance (ANOVA) evaluated task-specific differences in fatigue-induced asymmetry changes. Pearson correlation coefficients examined relationships between biomechanical variables. Multiple linear regression modeling identified independent predictors of asymmetry changes, incorporating age, BMI, disease duration, and WOMAC scores as independent variables.

Effect sizes were calculated using Cohen’s d for paired comparisons (21). Statistical significance was defined a priori at α = 0.05 (two-tailed). Assumptions for parametric testing (normality, homogeneity of variance) were verified using Shapiro-Wilk and Levene’s tests (22).

### Statistical Power and Sample Size

A priori power analysis (G*Power 3.1) determined that a sample size of *n* = 118 provided 95% statistical power to detect a medium effect size (Cohen’s d = 0.5) in paired comparisons of asymmetry indices between conditions, assuming a significance level of α = 0.05 and a correlation coefficient of 0.70 between repeated measures.

## Results

### Participant Characteristics

One hundred eighteen participants (mean age 62.6 ± 7.4 years, range 47–83 years) completed the study. The cohort comprised 62 males (52.5%) and 56 females (47.5%), with a mean BMI of 29.9 ± 4.2 kg/m^2^ (range 19.8–43.5 kg/m^2^). Mean disease duration was 8.2 ± 4.1 years, and mean WOMAC total score was 56.4 ± 18.2 (indicating moderate disease severity on average). No significant differences in demographic characteristics were observed between participants with right versus left knee involvement (*p* > 0.05 for all comparisons).

**Table 1.** Baseline participant characteristics (N = 118).

| Characteristic | Value | Range |
| --- | --- | --- |
| Sample size (n) | 118 | — |
| Age, years (mean ± SD) | 62.6 ± 7.4 | 47–83 |
| Sex, male / female | 62 (52.5%) / 56 (47.5%) | — |
| BMI, kg/m<sup>2</sup> (mean ± SD) | 29.9 ± 4.2 | 19.8–43.5 |
| Disease duration, years (mean ± SD) | 8.2 ± 4.1 | — |
| WOMAC total score (mean ± SD) | 56.4 ± 18.2 | 0–96 scale |

### Primary Outcome: Load Asymmetry Comparison

Paired t-test analysis revealed significantly increased inter-limb load asymmetry under fatigue conditions compared to non-fatigued baseline conditions (Fatigued: 34.2 ± 15.8%, Non-Fatigued: 25.5 ± 14.1%, mean difference 8.7 ± 12.3%, *t* (117) = 7.82, *p* < 0.001, Cohen’s d = 0.72). The 95% confidence interval for the mean difference was [6.3%, 11.1%].

**Figure 1.**
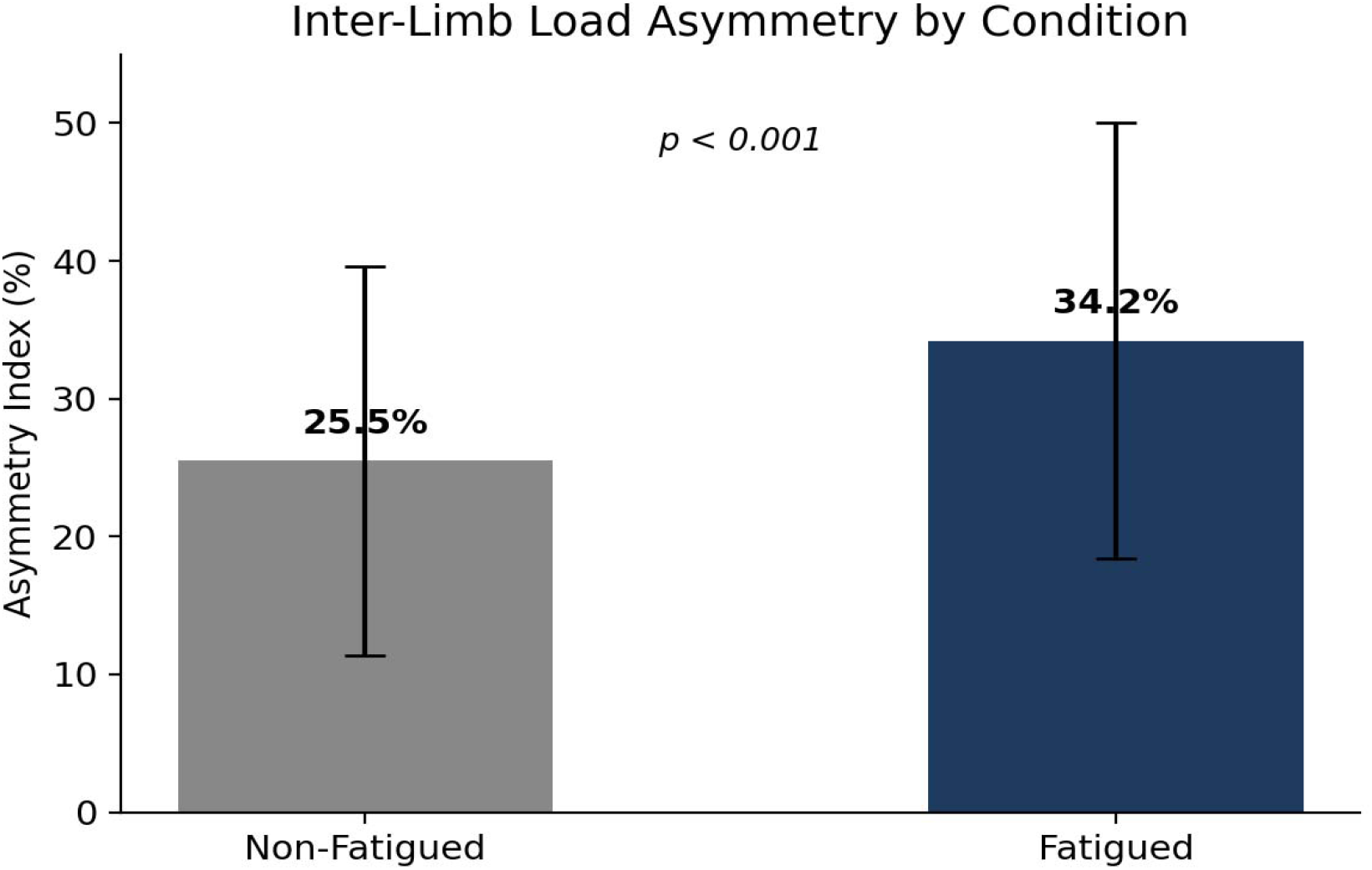
Mean inter-limb load asymmetry index under non-fatigued and fatigued conditions (error bars = SD).

### Task-Specific Analysis

One-way ANOVA revealed significant task-specific variations in fatigue-induced asymmetry changes (F (2,705) = 18.43, *p* < 0.001). Post-hoc pairwise comparisons (Tukey HSD) demonstrated:

- Stair negotiation: greatest asymmetry increase (11.2 ± 14.1%, *p* < 0.001)
- Sit-to-stand transitions: moderate increase (9.1 ± 11.8%, *p* < 0.001)
- Level walking: least pronounced change (5.3 ± 9.4%, *p* = 0.002)

These findings indicate that high-demand functional tasks demonstrate greater sensitivity to fatigue-related biomechanical alterations.

**Figure 2.**
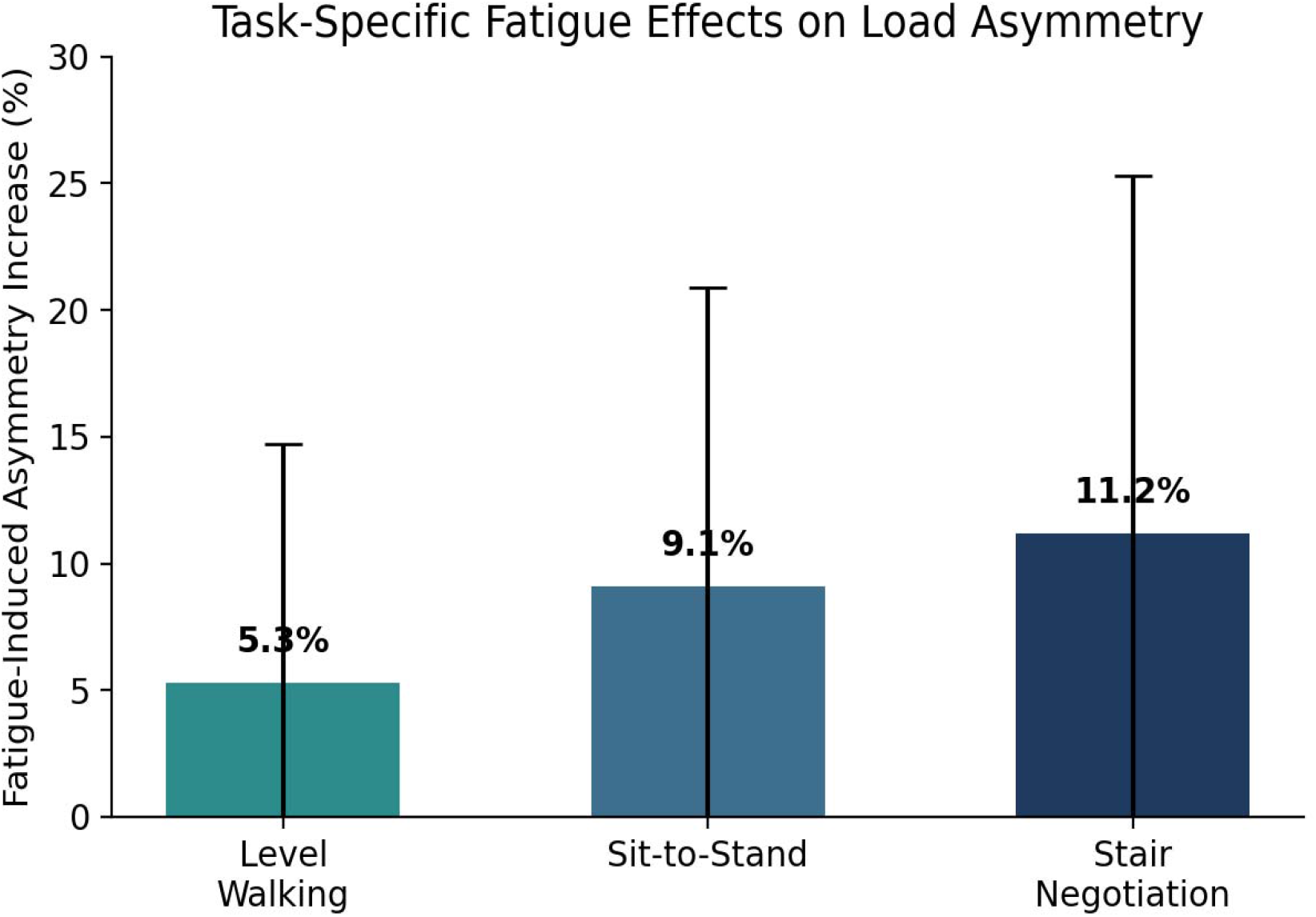
Fatigue-induced increase in inter-limb load asymmetry by functional task (error bars = SD).

### Loading Rate Correlations

Pearson correlation analysis in the fatigued condition revealed a strong inverse relationship between peak loading rate and time-to-peak load (*r* = -0.68, *p* < 0.001), suggesting that participants exhibiting delayed load application demonstrated reduced loading rates. This relationship was weaker in the non-fatigued condition (*r* = -0.42, *p* < 0.001), indicating condition-dependent changes in neuromuscular control.

**Figure 3.**
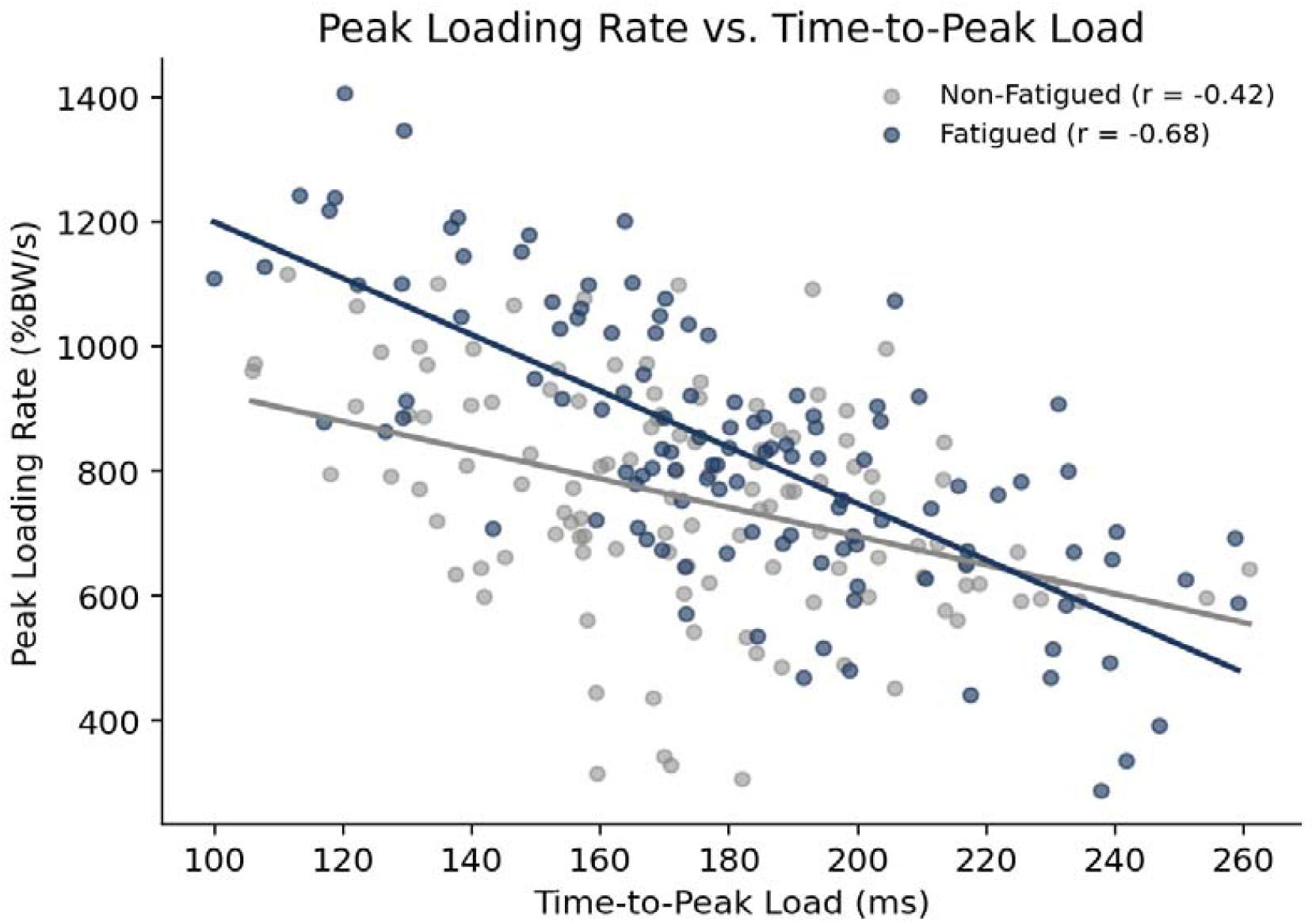
Relationship between peak loading rate and time-to-peak load under non-fatigued and fatigued conditions.

### Predictors of Asymmetry Response to Fatigue

Multiple linear regression modeling identified significant independent predictors of asymmetry change:

### Model Summary

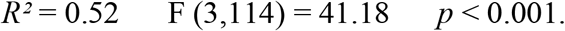

### Predictors

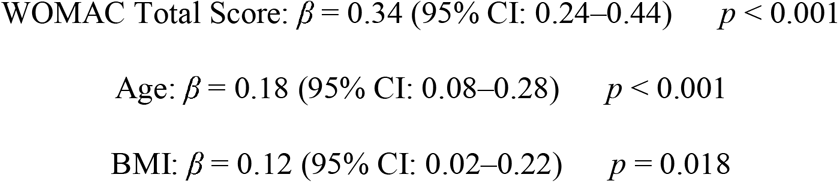

The model indicates that greater disease severity, older age, and higher BMI were associated with more pronounced fatigue-induced increases in load asymmetry. Disease duration was not a significant independent predictor (p = 0.156) after accounting for other variables.

### Peak Load Distribution

Analysis of peak vertical ground reaction forces on the affected knee revealed minimal change between conditions (Non-Fatigued: 87.3 ± 18.2 % BW, Fatigued: 86.8 ± 19.1 % BW, *p* = 0.412). In contrast, the unaffected knee demonstrated a significant increase in loading under fatigue conditions (Non-Fatigued: 112.7 ± 16.4 % BW, Fatigued: 121.4 ± 17.8 % BW, *p* < 0.001), indicating that fatigue-induced asymmetry primarily resulted from increased loading on the unaffected limb rather than decreased loading on the affected limb.

**Figure 4.**
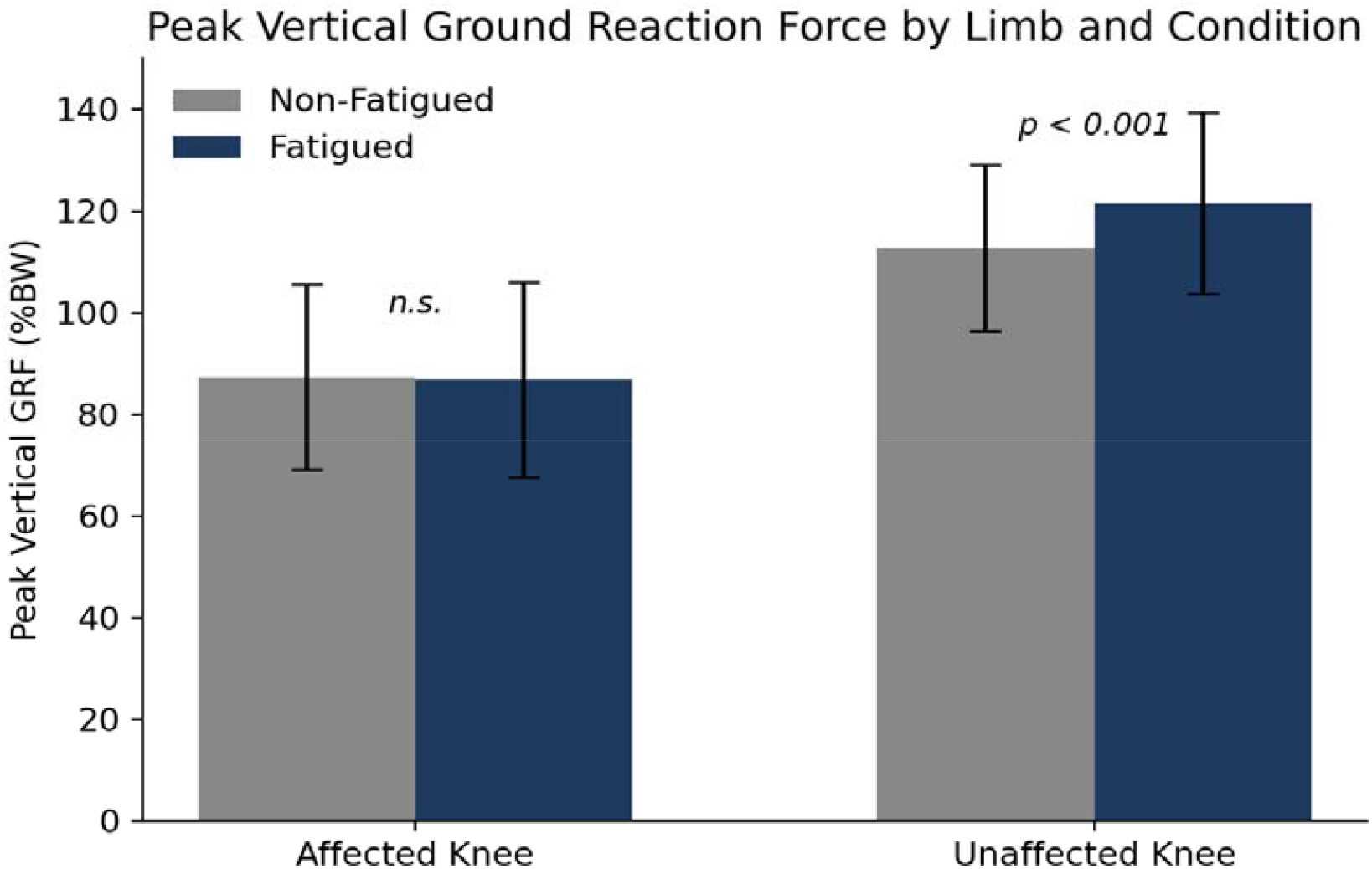
Peak vertical ground reaction force (%BW) on the affected and unaffected knee under non-fatigued and fatigued conditions.

**Table 2.**
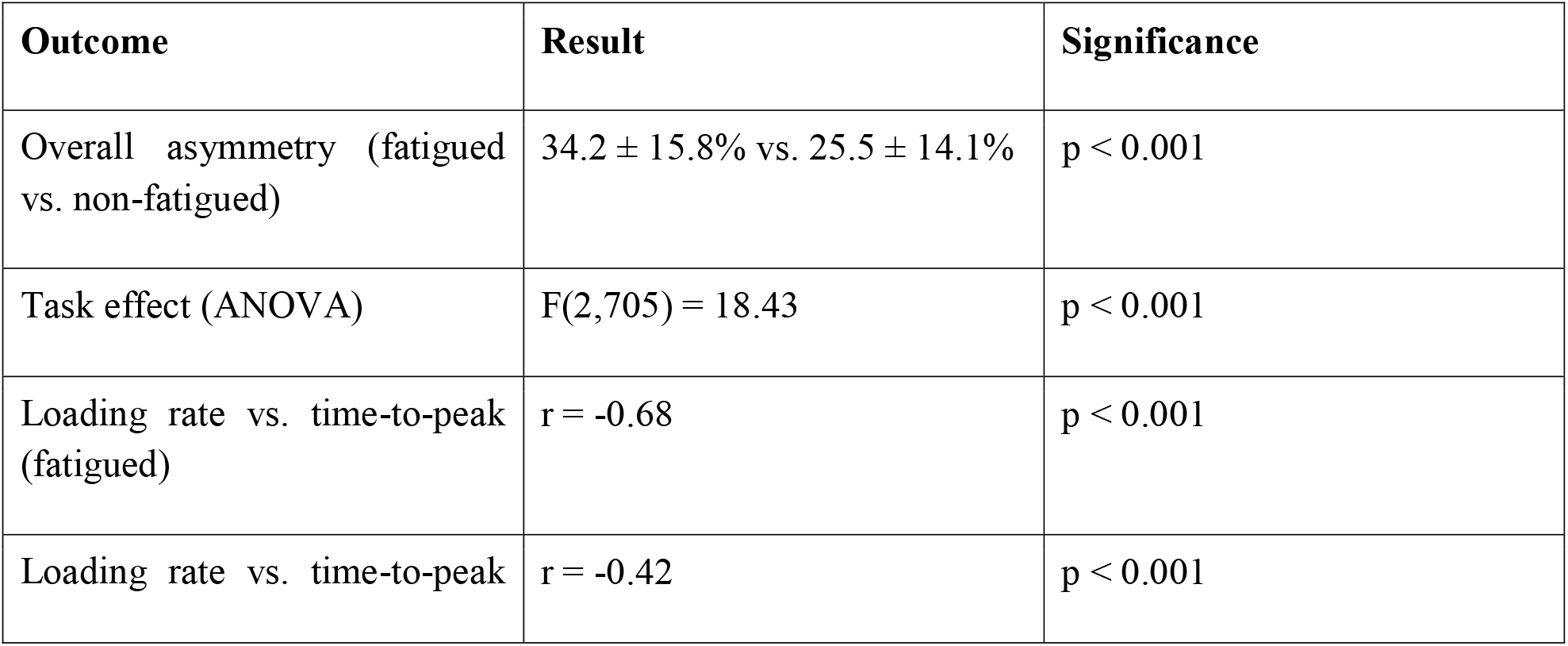
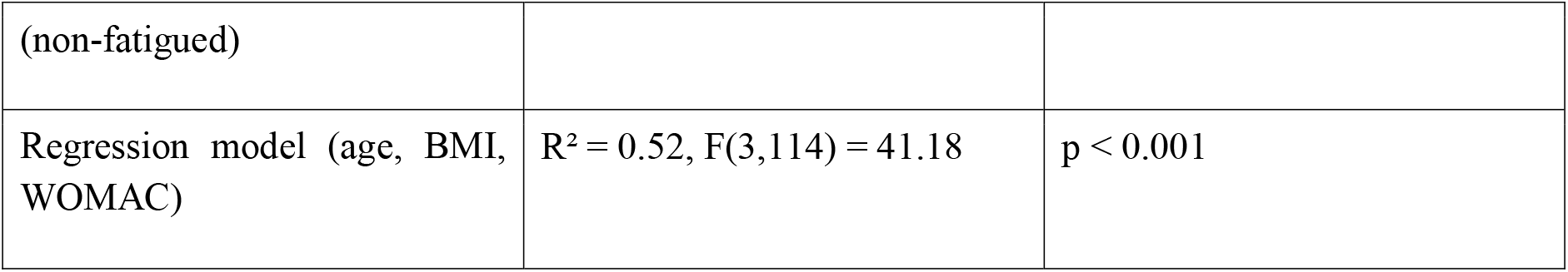
Summary of key statistical findings.

## Discussion

This study presents empirical evidence demonstrating that physical fatigue significantly exacerbates inter-limb load asymmetry in individuals with unilateral knee OA, with pronounced effects during high-demand functional tasks. The mean 8.7% increase in asymmetry index represents a clinically meaningful deterioration in load distribution, as longitudinal evidence suggests that sustained asymmetries of this magnitude are associated with accelerated radiographic OA progression (23, 24).

The task-specific pattern observed in this study aligns with biomechanical theory regarding neuromuscular demand and fatigue susceptibility. Stair negotiation, which demands the greatest muscular effort and generates the highest joint reaction forces, demonstrated the most pronounced fatigue-induced asymmetry changes (12, 25). Conversely, level walking, representing a more automatic motor task with lower metabolic demand, showed relatively attenuated asymmetry changes, consistent with prior work showing that increased knee joint loading during walking is itself associated with OA severity (26).

The mechanistic underpinning of observed fatigue effects likely involves fatigue-induced alterations in neuromuscular control. Acute fatigue is known to reduce motor unit recruitment amplitude, increase motor unit synchronization, and impair proprioceptive feedback mechanisms (27), fatigue has similarly been shown to blunt peripheral sensory acuity in other body regions, suggesting a generalized effect on afferent feedback under conditions of muscular fatigue (28). In OA populations, these changes may disproportionately affect the affected limb, which has baseline muscular deficits due to pain-related inhibition and chronic disuse (29). This is consistent with evidence that unilateral knee OA is accompanied by contralateral limb strength deficits and impaired dynamic postural control (30), and that knee extensor and flexor strength independently predict joint biomechanics during functional tasks (31).

These findings carry direct implications for rehabilitation strategy. Because muscular strength appears protective against both the onset and progression of lower-limb OA, including hip abduction moment reductions in medial tibiofemoral disease and thigh muscle strength in incident hip OA (32), targeted strengthening programs may help offset fatigue-induced asymmetry. Structured exercise interventions, including land-based programs shown to be effective for knee OA in systematic review (33), water-based exercise therapy (34), and combined exercise-and-weight-loss programs such as the Arthritis, Diet, and Activity Promotion Trial (35), represent viable avenues for reducing fatigue-related biomechanical deterioration. Home-based exercise programs have also been shown to reduce fall risk in older adults with impaired gait control, a population that shares mechanistic overlap with fatigued OA patients (36). Broader pain-management modalities, such as acupuncture for chronic musculoskeletal pain (37), and general principles of muscular joint stabilization drawn from the lumbar spine literature (38, 39), may further inform multidisciplinary management of fatigue-related load asymmetry in knee OA.

Clinical management strategies should incorporate fatigue mitigation alongside traditional exercise-based interventions. Future investigations should employ longitudinal designs with radiographic assessments to determine whether fatigue-related acute biomechanical changes predict structural disease progression.

## Conclusions

This investigation demonstrates that physical fatigue significantly increases inter-limb load asymmetry in unilateral knee OA, with task-specific effects greatest during high-demand functional activities such as stair negotiation. Disease severity, age, and BMI independently predict the magnitude of fatigue-induced asymmetry changes. These findings suggest that fatigue represents a modifiable risk factor for biomechanical deterioration in OA populations. Clinical management strategies should incorporate fatigue mitigation alongside traditional exercise-based interventions.

## Data Availability

All data produced in the present study are available upon reasonable request to the corresponding author.

## Notes

**Conflict of Interest:** The authors have declared no conflict of interest.

### Competing Interest Statement

The authors have declared no competing interest.

### Author Declarations

Ethics committee of Institutional Review Board of Islam College of Physical Therapy gave ethical approval for this work.

